# Risk factors for arterial stiffness in a multi-ethnic Asian population: Results from the Health for Life in Singapore Study

**DOI:** 10.1101/2025.10.10.25337779

**Authors:** Harinakshi Sanikini, Theresia Mina, Konstantinos K Tsilidis, The HELIOS Study Team, Joanne Ngeow, Eng Sing Lee, Jimmy Lee, Paul Elliott, John C Chambers, Elio Riboli

## Abstract

**Background:** Arterial stiffness (AS) measured by pulse wave velocity (PWV) is a well-recognized marker for increased cardiovascular disease risk in Asian and Western individuals. We evaluated factors in relation to AS using the exposure-wide association study (EWAS) approach within the Health for Life in Singapore (HELIOS) study. Subsequently, two-sample Mendelian randomization (MR) analysis was utilized to examine potential causal associations between identified factors and AS.

**Methods:** The HELIOS study is a multi-ethnic prospective cohort study that included adults aged 30-85 years. We analyzed 8,880 subjects of Chinese, Malay, and South Asian ethnicity participating in the HELIOS cohort. PWV was measured by Vicorder. We assessed 54 predictor variables using EWAS approach and correction for multiple comparisons was applied using the false discovery rate (FDR < 5%) method. Linear regression models were used to assess the association of each predictor variable with PWV. Inverse-variance-weighted MR method was used to estimate causal effects.

**Results:** Of the 54 variables, 41 variables were significantly associated with PWV at FDR <5%. We found age, systolic blood pressure (SBP), diastolic blood pressure, and anthropometric, and body composition measurements were positively associated with PWV. Furthermore, insulin, glycated hemoglobin (HbA1c), glucose, triglycerides, albumin, alanine transaminase, gamma-glutamyl transferase, C-reactive protein, and uric acid were positively associated with PWV. Inverse associations were observed for PWV with high-density lipoprotein cholesterol (HDL-C), potassium, vitamin D, bilirubin, handgrip strength and educational level. These associations appear stronger among Chinese participants than among Malay and South Asian participants. In MR analyses, genetically predicted SBP was positively with AS, while genetically predicted HDL-C and HbA1c were inversely associated with AS.

**Conclusions:** Our systematic evaluation provides new knowledge on the complex array of anthropometric, body composition, physiological, biochemical, behavioral, and sociodemographic correlates of PWV. Our MR analysis indicates a potential causal association between SBP, HDL-C, HbA1c and AS.

## Introduction

Cardiovascular diseases, predominantly driven by ischemic heart disease and stroke, remain the leading cause of death and disability worldwide^1^. The global distribution for cardiovascular disease has been progressively shifting away from Europe and North America, and towards emerging market economies of the Asia-Pacific region. The World Health Organization report that the proportion of deaths caused by cardiovascular disease in Asia-Pacific region has increased from 23% in 1990 to 35% in 2019^2^. Deaths from cardiovascular disease are also more likely to occur prematurely (<70 years of age) in Asia, compared to Europe and the US (39% vs 22% and 23% respectively, 2019)^2^. Patterns of cardiovascular disease show important variation between the global regions^2^. Whilst ischemic heart disease is the major cause of cardiovascular deaths in Central, Southern and Western Asia, death from stroke is most common in East and Southeast Asia in 2019^2^. There is an urgent need to better identify Asian individuals who are at high risk of cardiovascular disease, and to better understand the reasons for differing patterns of cardiovascular disease between population groups, to enable national and global efforts aimed at improving health and human potential.

Arterial stiffness is a well-recognized marker for increased cardiovascular disease risk, including myocardial infarction, heart failure, and total mortality, as well as stroke, dementia, and renal disease^3^. Arterial stiffness arises through the loss of elasticity in vascular wall, reflecting dysregulation in the turnover of collagen, elastin, and other vascular scaffolding protein, as well as changes in extracellular matrix, smooth muscle biology and extrinsic factors such as neuro-hormonal signalling^3^. Arterial stiffness increases with age, and is closely associated with diabetes, hypertension, physical inactivity, and cigarette smoking^4^.

Although epidemiological studies of arterial stiffness have predominantly been carried out in European and North American populations, available evidence suggests that arterial stiffness mostly measured by pulse wave velocity (PWV) is also a marker of cardiovascular risk in Asian individuals^5^. Diabetes, hypertension, body mass index (BMI), circulating markers of lipid metabolism, liver and renal function, and systemic inflammation have been positively associated with arterial stiffness in studies of East Asian populations^6–11^. However, previous studies investigating the causal associations between identified risk factors and arterial stiffness are limited^12,13^.

The aim of this study was to evaluate an extensive list of factors (anthropometry measurements, body composition, biochemical and physiological parameters, sociodemographic factors, and lifestyle behaviors) in relation to arterial stiffness using the exposure-wide association study (EWAS) approach within the Health for Life in Singapore (HELIOS) study. In place of testing one or few associations at a time, an EWAS approach follows methodological principles similar to the genome-wide association studies (GWAS), which involves the assessment of various factors for associations with health outcome using multiple comparison adjustments^14^. Furthermore, we examined the potential causal associations between identified factors and arterial stiffness using two-sample Mendelian randomization (MR) approach.

## Methods

### Study population

The HELIOS study is an ongoing multi-ethnic prospective cohort study aimed at investigating the associations between diet, lifestyle, genetic and environmental factors, and the development of chronic diseases in Singapore^15^. Approximately, 10,000 Singaporean men and women, aged 30-85 years were recruited between 2018 and 2022 from the general population through various community outreach initiatives to ensure representation across ethnic and socio-economic groups. Ethnicity was self-reported and categorized into three groups: Chinese, Malay, and South Asian (India and from other countries in the Indian subcontinent). Ethical approval of the HELIOS study was obtained from the Nanyang Technological University’s Institutional Review Board (IRB-2016-11-030). All participants signed informed consent form.

For this study, we excluded participants with missing data for the outcomes, exposures, confounders, and extreme PWV outliers, leaving 8,880 participants available for analysis.

### Predictor variables

*<u>Anthropometry and body composition measurements</u>*: Body weight and height were measured using computerized measuring instruments^15^. A non-stretchable sprung tape measure was used to measure waist circumference (WC) and hip circumference (HC). BMI was calculated as weight in kilograms divided by height in meters squared, and waist-to-hip ratio (WHR) computed as WC (cm) divided by HC (cm). The Tanita BC-418 body composition analyzer was used to assess body composition measures including body fat, muscle mass and fat mass.

*<u>Blood pressure:</u>* The blood pressure (systolic blood pressure (SBP)/diastolic blood pressure (DBP)) was measured three times in the right arm using an Omron blood pressure monitor^15^. We used the mean of the three SBP and DBP measurements.

*<u>Biochemical measurements</u>:* A 60 mL blood sample was collected for participants at the time of recruitment^15^. Blood samples were processed according to standardized protocol and aliquots were stored in small cryovials (−80 °C freezers). A panel of biomarkers was measured in blood samples using ADVIA 1800 chemistry system (Siemens Healthcare, Munich, Germany). The measured biomarkers included: 1) lipid profile 2) insulin, glucose, and glycated hemoglobin A1c (HbA1c) 3) renal function test (creatinine, uric acid, urea) 4) liver function test (albumin, bilirubin, alanine transaminase (ALT), gamma-glutamyl transferase (GGT)).

*Sociodemographic factors/Lifestyle factors:* A computerized self-administered questionnaire was used to collect information on sociodemographic characteristics, lifestyle factors, health, and medical history^15^.

*Handgrip strength:* The Jamar Plus Hand Dynamometer was used to measure right and left handgrip strength^15^.

### Outcome

*<u>Arterial stiffness:</u>* The Vicorder was used to measure PWV^15^. Participants were asked to lie down at a 30-degree angle on a couch, where blood pressure cuffs were placed over the brachial artery on the right upper arm and over the femoral artery on the right thigh. The pressure within the cuffs increases and fluctuations around this pressure was used to determine a pulse waveform that is closely related to intra-arterial pressure.

### Statistical analysis

PWV was classified into tertiles <11, 11-14, >14 (m/sec). Median and interquartile range (25^th^ and 75^th^ percentile) or frequencies were computed for baseline characteristics of total participants by gender, ethnic group, and based on PWV categories.

We used EWAS approach to examine the cross-sectional associations of a large array of potential correlations with PWV. The evaluated predictors of PWV included 54 variables, which were categorized to anthropometry (height, weight, BMI, WC, HC, and WHR), blood pressure (SBP, and DBP), body composition (body fat mass (BFM), fat free mass (FFM), skeletal muscle mass (SMM), PBF (percent body fat), and visceral fat level (VFL)), biochemical (total cholesterol (TC), high-density lipoprotein cholesterol (HDL-C), low-density lipoprotein cholesterol (LDL-C), triglycerides, TC to HDL-C ratio (TC/HDL-C), insulin, HbA1c, glucose, albumin, bilirubin, ALT, GGT, C-reactive protein (CRP), creatinine, uric acid, urea, potassium, and vitamin D), handgrip strength (right and left), sociodemographic factors (age {<35, 35-39, 40-44, 49-45, 50-54, 55-59, ≥60 years, ethnicity {Chinese, Malay, and South Asian}, educational level {none, primary/equivalent, secondary/equivalent, junior college/vocational/diploma/equivalent, and university}), and lifestyle behaviors (smoking status {never, former and current}, physical activity {low, moderate and high}, and alcohol intake frequency {never, special occasions only, 1-3 times/month, 1-2 times/week, 3-4 times a week, and daily or mostly}).

All predictor variables were log-transformed to normalize their distribution, and they were standardized and included as continuous variables in the statistical models to reflect associations per one standard deviation (SD) increase. For sociodemographic and lifestyle variables, dummy variables were created where the lowest category was used as a reference group. Separate linear regression models were used to assess the association of each predictor variable with PWV, adjusting for age, gender and ethnicity as covariates. We corrected for multiple comparisons by estimating an adjusted p value (q value) using the false discovery rate (FDR) approach^14^. Subgroup analyses were performed for gender and ethnicity. Sensitivity analyses were performed by excluding participants in whom diabetes was self-reported and participants with hypertension (self-reported or SBP/DBP at recruitment ≥140/90 mmHg). A heatmap with Pearson correlation coefficients adjusted for age, gender, and ethnicity was plotted for the predictor variables. To determine independent effects among variables, we fitted multivariable models for PWV in all participants and by ethnic group, separately considering all the FDR significant variables. Multicollinearity was assessed by correlation matrices (r >0.7, and variance inflation factor >4). All analyses were performed in R version 4.4 and SAS 9.4 software.

#### MR assumptions

To perform MR, three assumptions must be satisfied about the genetic variants^16^: 1) it should be associated with exposure; 2) it should not associated with any confounders between the exposure and outcome and 3) it should be related to the outcome only through the exposures. In this study, we conducted a two-sample MR approach using GWAS summary statistics to examine the potential causal associations between predictor variables and PWV, specifically on those that were independently linked to PWV in multivariable analysis.

#### Source of genetic instruments

All the GWAS summary statistics are publicly available and were sourced from GWAS Catalog (https://www.ebi.ac.uk/gwas/). Details of GWAS studies used to select genetic instruments for both exposures and outcome are presented in Table 1. Briefly, summary statistics data for GWAS on predictor variables were obtained from the International Consortium for Blood Pressure (ICBP) for SBP^17^, Meta-Analyses of Glucose and Insulin-related traits Consortium (MAGIC) for HbA1c^18^, Global Lipids Genetics Consortium (GLGC) for HDL-C^19^, a combined analysis of Biobank Japan, UK Biobank and Finn Gen for albumin^20^, Social Science Genetic Association Consortium (SSGAC) for educational level^21^, UK Biobank study for potassium^22^, and handgrip strength^23^, respectively. Summary-level genetic data for arterial stiffness (outcome) was obtained from the UK Biobank study, where arterial stiffness was measured by arterial stiffness index^24^. Details of all these studies including study design and methods, genotyping, imputation, and quality control have been described elsewhere^17–24^.

**Table 1.**
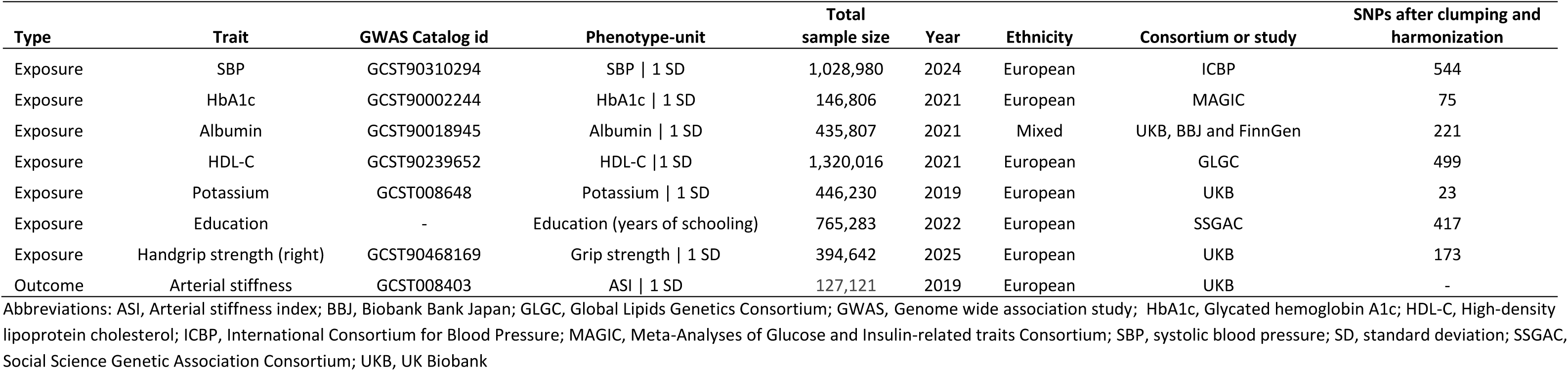
Summary of GWAS studies used in this study.

| Type | Trait | GWAS Catalog id | Phenotype-unit | Total sample size | Year | Ethnicity | Consortium or study | SNPs after clumping and harmonization |
| --- | --- | --- | --- | --- | --- | --- | --- | --- |
| Exposure | SBP | GCST90310294 | SBP \| 1 SD | 1,028,980 | 2024 | European | ICBP | 544 |
| Exposure | HbA1c | GCST90002244 | HbA1c \| 1 SD | 146,806 | 2021 | European | MAGIC | 75 |
| Exposure | Albumin | GCST90018945 | Albumin \| 1 SD | 435,807 | 2021 | Mixed | UKB, BBJ and FinnGen | 221 |
| Exposure | HDL-C | GCST90239652 | HDL-C \| 1 SD | 1,320,016 | 2021 | European | GLGC | 499 |
| Exposure | Potassium | GCST008648 | Potassium \| 1 SD | 446,230 | 2019 | European | UKB | 23 |
| Exposure | Education | - | Education (years of schooling) | 765,283 | 2022 | European | SSGAC | 417 |
| Exposure | Handgrip strength (right) | GCST90468169 | Grip strength \| 1 SD | 394,642 | 2025 | European | UKB | 173 |
| Outcome | Arterial stiffness | GCST008403 | ASI \| 1 SD | 127,121 | 2019 | European | UKB | - |

Abbreviations: ASI, Arterial stiffness index; BBJ, Biobank Bank Japan; GLGC, Global Lipids Genetics Consortium; GWAS, Genome wide association study; HbA1c, Glycated hemoglobin A1c; HDL-C, High-density lipoprotein cholesterol; ICBP, International Consortium for Blood Pressure; MAGIC, Meta-Analyses of Glucose and Insulin-related traits Consortium; SBP, systolic blood pressure; SD, standard deviation; SSGAC, Social Science Genetic Association Consortium; UKB, UK Biobank

#### Selection of instrumental variables and MR analysis

We selected single nucleotide polymorphisms (SNPs) as instruments based on their association with exposure of interest at genome-wide significance (p < 5 ×10^-8^). To retain only independent SNPs, those in linkage disequilibrium were excluded through clumping using an r² threshold of 0.001 and a clumping distance of 10 kilobases. Genetic instruments for each predictor are shown in Table S1 to S7.

For MR analysis, random effect inverse-variance weighted (IVW) model was used as the primary MR method to assess associations of genetically predicted SBP, HbA1c, HDL-C, albumin, potassium, educational level, and handgrip strength with arterial stiffness. These predictor variables were identified through multivariable analysis. The IVW method provides unbiased estimate, under the assumption that genetic variants are uncorrelated and independent^16^. Sensitivity analyses were performed using other MR methods, including MR-Egger that assess pleiotropic effects of instrumental variables, and the weighted-median approach, which assumes at least 50% of weight in the analysis comes from instrumental variables^16^. Additionally, to assess the direct effects of lipid traits on arterial stiffness, multivariable MR (MVMR) analyses were conducted using summary statistics from the GLGC^19^, with HDL-C, LDL-C and triglycerides included in the model. All analyses were performed with R version (4.4) using the ieugwasr, MendelianRandomization, TwoSampleMR and MVMR packages.

## Results

### Characteristics of the HELIOS study participants

Of the total participants (59.3% women), 68.2% were Chinese, 13.5% were Malay, and 18.2% were South Asian. Baseline characteristics of total participants and by ethnic group are shown in Table 2. Men had higher WC, SBP, DBP, lower PBF, higher ALT and creatinine levels, and higher handgrip strength and they were highly educated, more physically active, more likely to smoke, and drink alcohol compared to women. Furthermore, the prevalence of self-reported diabetes is higher in men than in women. Among ethnic groups (Table 2), Chinese and South Asian participants were slightly older than Malay participants. Chinese participants also had lower BMI, WC, PBF, and CRP levels and they were highly educated, less likely to smoke, and less physically active compared to Malay and South Asian participants. Furthermore, South Asian participants had slightly higher creatinine levels than Chinese and Malay participants, while Malay participants was less likely to drink alcohol than Chinese and South Asian participants. The prevalence of self-reported diabetes was lower in Chinese participants than in Malay and South Asian participants.

**Table 2.** Baseline characteristics of all the participants by ethnicity in the HELIOS study.

|  | All |  |  | Chinese |  |  | Malay |  |  | South Asian |  |  |
| --- | --- | --- | --- | --- | --- | --- | --- | --- | --- | --- | --- | --- |
|  | All | Men | Women | All | Men | Women | All | Men | Women | All | Men | Women |
| **Participants (n)** | 8880 | 3617 | 5263 | 6062 | 2408 | 3654 | 1200 | 421 | 779 | 1618 | 788 | 830 |
| **Age\* (years)** | 53.1<br>(43.1-61.4) | 52.7<br>(41.7-62.0) | 53.3<br>(44.2-61.0) | 53.4<br>(43.3-62.2) | 53.1<br>(42.1-62.8) | 53.7<br>(44.3-61.9) | 51.5<br>(41.4-58.1) | 50.4<br>(39.3-58.4) | 51.8<br>(43.1-58.0) | 53.3<br>(43.9-60.7) | 53.2<br>(42.9-61.0) | 53.5<br>(44.8-60.4) |
| **BMI\*(kg/m<sup>2</sup>)** | 24.1<br>(21.6-27.3) | 24.8<br>(22.6-27.5) | 23.5<br>(21.0-27.0) | 23.1<br>(21.0-25.6) | 24.0<br>(22.1-26.4) | 22.4<br>(20.4-24.9) | 27.5<br>(24.5-31.3) | 27.5<br>(24.7-31.2) | 27.5<br>(24.3-31.4) | 26.4<br>(23.7-29.5) | 26.1<br>(23.7-28.7) | 26.7<br>(23.7-30.3) |
| **WC\* (cm)** | 82.2<br>(74.6-90.2) | 87.0<br>(80.9-94.6) | 78.1<br>(71.5-86.0) | 79.8<br>(73.0-87.0) | 85.0<br>(79.6-91.5) | 35.0<br>(30.4-39.1) | 86.8<br>(80.0-95.3) | 91.4<br>(84.1-99.8) | 84.8<br>(77.6-92.0) | 89.1<br>(81.3-96.8) | 92.3<br>(85.1-99.4) | 86.0<br>(78.3-93.8) |
| **PBF\* (%)** | 33.2<br>(26.8-39.2) | 27.0<br>(22.7-32.2) | 36.9<br>(32.2-42.1) | 31.4<br>(25.6-36.7) | 25.6<br>(21.5-30.0) | 35.0<br>(30.4-39.1) | 39.2<br>(32.7-44.2) | 31.8<br>(25.9-36.0) | 42.3<br>(37.8-46.5) | 36.7<br>(29.5-43.5) | 30.1<br>(25.6-35.2) | 42.7<br>(38.0-47.3) |
| **SBP\* (mmHg)** | 118 (108-131) | 122 (133-133) | 116 (104-130) | 118 (108-131) | 122 (113-133) | 115 (104-129) | 119 (110-132) | 122 (113-134) | 118 (107-131) | 120 (109-132) | 121 (112-132) | 117 (107-132) |
| **DBP\* (mmHg)** | 74.7<br>(67.7-82.3) | 78.3<br>(72.3-85.7) | 72.0<br>(65.3-79.3) | 74.7<br>(67.3-82.0) | 78.3<br>(72.3-85.3) | 71.3<br>(65.0-79.0) | 74.6<br>(67.7-82.3) | 78.3<br>(72.0-85.7) | 72.7<br>(66.0-80.0) | 76.0<br>(69.3-83.0) | 78.7<br>(72.7-86.0) | 72.7<br>(66.7-80.3) |
| **HDL-C\* (mmol/L)** | 1.5 (1.2-1.8) | 1.3 (1.1-1.5) | 1.6 (1.4-1.9) | 1.6 (1.3-1.9) | 1.4 (1.2-1.6) | 1.7 (1.4-2.0) | 1.4 (1.2-1.6) | 1.2 (1.1-1.4) | 1.5 (1.3-1.7) | 1.2 (1.1-1.5) | 1.1 (1.0-1.8) | 1.4 (1.2-1.6) |
| **LDL-C\* (mmol/L)** | 3.1 (2.5-3.7) | 3.1 (2.5-3.7) | 3.1 (2.6-3.6) | 3.0 (2.5-3.6) | 3.1 (2.5-3.6) | 3.0 (2.6-3.6) | 3.2 (2.6-3.8) | 3.3 (2.7-4.0) | 3.1 (2.6-3.8) | 3.1 (2.5-3.7) | 3.1 (2.5-3.7) | 3.1 (2.6-3.6) |
| **Triglycerides\* (mmol/L)** | 1.1 (0.8-1.5) | 1.2 (0.9-1.7) | 1.0 (0.7-1.4) | 1.0 (0.8-1.4) | 1.2 (0.8-1.6) | 1.0 (0.7-1.3) | 1.2 (0.9-1.7) | 1.4 (1.0-2.0) | 1.1 (0.8-1.5) | 1.2 (0.9-1.6) | 1.3 (1.0-1.8) | 1.1 (0.9-1.5) |
| **Glucose\* (mmol/L)** | 4.8 (4.5-5.2) | 4.9 (4.6-5.3) | 4.7 (4.5-5.1) | 4.8 (4.5-5.1) | 4.9 (4.6-5.2) | 4.7 (4.5-5.0) | 4.8 (4.5-5.3) | 4.9 (4.6-5.5) | 4.8 (4.5-5.3) | 4.9 (4.6-5.5) | 5.0 (4.7-5.8) | 4.9 (4.5-5.3) |
| **HbA1c\* (mmol/L)** | 5.5 (5.3-5.8) | 5.6 (5.3-5.9) | 5.5 (5.3-5.8) | 5.5 (5.3-5.8) | 5.5 (5.3-5.8) | 5.5 (5.2-5.8) | 5.6 (5.4-6.0) | 5.7 (5.4-6.1) | 5.6 (5.3-5.9) | 5.7 (5.4-6.1) | 5.7 (5.4-6.2) | 5.6 (5.4-6.0) |
| **ALT\* (U/L)** | 21.2<br>(16.1-29.4) | 26.3<br>(20.0-36.7) | 18.4<br>(14.6-24.3) | 20.9<br>(15.9-28.6) | 25.6<br>(19.7-34.9) | 18.4<br>(14.5-24.1) | 21.3<br>(16.2-31.4) | 29.2<br>(20.2-42.5) | 19.0<br>(14.8-25.6) | 22.3<br>(16.5-31.1) | 27.6<br>(21.0-38.1) | 18.2<br>(14.8-24.3) |
| **Creatinine\* (umol/L)** | 66.0<br>(57.0-79.0) | 81.0<br>(75.0-89.0) | 58.0<br>(53.0-64.0) | 65.0<br>(56.0-79.0) | 81.0<br>(75.0-89.0) | 58.0<br>(53.0-64.0) | 65.0<br>(57.0-78.0) | 83.0<br>(75.0-91.0) | 59.0<br>(54.0-66.0) | 69.0<br>(58.0-82.0) | 81.0<br>(74.0-90.0) | 59.0<br>(54.0-66.0) |
| **CRP\* (mg/L)** | 0.4 (0.1-1.6) | 0.4 (0.1-1.2) | 0.5 (0.1-1.9) | 0.3 (0.1-1.0) | 0.3 (0.1-0.9) | 0.3 (0.1-1.1) | 1.1 (0.3-3.2) | 0.7 (0.3-2.4) | 1.3 (0.3-3.6) | 1.2 (0.4-3.7) | 0.8 (0.3-2.2) | 2.2 (0.6-5.2) |
| **Handgrip strength (right)\*** | 27.0<br>(21.7-34.8) | 36.5<br>(31.1-42.0) | 23.0<br>(19.7-26.7) | 27.1<br>(21.9-34.9) | 36.7<br>(31.5-42.0) | 23.2<br>(19.9-26.9) | 25.6<br>(20.9-33.0) | 36.1<br>(30.2-42.3) | 22.7<br>(19.3-26.4) | 27.7<br>(21.9-35.7) | 35.7<br>(30.5-41.5) | 22.6<br>(19.1-26.0) |
| **Education level (%)** |  |  |  |  |  |  |  |  |  |  |  |  |
| None/primary school not completed | 0.9 | 0.5 | 1.1 | 0.8 | 0.5 | 1.0 | 1.8 | 0.5 | 2.6 | 0.5 | 0.4 | 0.6 |
| Primary/equivalent | 3.1 | 2.1 | 3.8 | 2.4 | 1.9 | 2.8 | 5.2 | 3.3 | 6.2 | 4.0 | 2.0 | 5.8 |
| Secondary school/equivalent | 20.4 | 14.6 | 24.4 | 17.0 | 10.5 | 21.4 | 32.0 | 24.9 | 35.8 | 24.6 | 21.7 | 27.4 |
| Junior college/vocational/diploma/<br>equivalent | 29.5 | 29.9 | 29.1 | 28.2 | 27.7 | 28.5 | 38.8 | 45.4 | 35.2 | 27.3 | 28.6 | 26.1 |
| University | 45.5 | 52.2 | 40.9 | 50.9 | 58.8 | 45.8 | 21.7 | 24.9 | 19.9 | 42.8 | 46.7 | 39.2 |
| Missing | 0.7 | 0.8 | 0.6 | 0.6 | 0.8 | 0.6 | 0.6 | 1.0 | 0.4 | 0.8 | 0.6 | 1.0 |
| **Smoking status (%)** |  |  |  |  |  |  |  |  |  |  |  |  |
| Never | 82.1 | 68.2 | 91.6 | 85.7 | 74.3 | 93.1 | 67.9 | 40.6 | 82.7 | 79.2 | 64.0 | 93.6 |
| Former | 8.7 | 16.3 | 3.5 | 7.6 | 14.3 | 3.1 | 13.8 | 26.8 | 6.8 | 9.4 | 16.8 | 2.4 |
| Current | 8.3 | 14.8 | 3.8 | 5.8 | 10.6 | 2.7 | 17.5 | 31.8 | 9.8 | 10.6 | 18.7 | 2.9 |
| Missing | 0.9 | 0.7 | 1.1 | 1.0 | 0.8 | 1.1 | 0.8 | 0.7 | 0.8 | 0.9 | 0.6 | 1.1 |
| **Physical activity (%)** |  |  |  |  |  |  |  |  |  |  |  |  |
| Low | 15.9 | 16.0 | 15.9 | 15.3 | 14.2 | 16.0 | 18.3 | 20.2 | 17.3 | 16.3 | 18.9 | 13.9 |
| Moderate | 46.3 | 41.2 | 49.9 | 45.5 | 41.4 | 48.3 | 46.2 | 38.7 | 50.2 | 49.5 | 42.0 | 56.6 |
| High | 21.9 | 28.3 | 17.5 | 19.8 | 26.0 | 15.7 | 26.3 | 33.5 | 22.5 | 26.6 | 32.7 | 20.8 |
| Missing | 15.8 | 14.5 | 16.7 | 19.4 | 18.4 | 20.0 | 9.2 | 7.6 | 10.0 | 7.5 | 6.4 | 8.7 |
| **Alcohol intake frequency (%)** |  |  |  |  |  |  |  |  |  |  |  |  |
| Never | 40.9 | 32.2 | 46.9 | 28.9 | 20.7 | 34.3 | 91.1 | 90.0 | 91.7 | 48.6 | 36.3 | 60.2 |
| Special occasions only | 38.3 | 36.4 | 39.6 | 48.0 | 45.6 | 49.5 | 5.5 | 5.0 | 5.8 | 26.4 | 24.8 | 28.0 |
| 1-3 times/month | 8.9 | 12.7 | 6.4 | 10.4 | 14.2 | 7.8 | 1.0 | 1.4 | 0.8 | 9.5 | 14.0 | 5.2 |
| 1-2 times/week | 6.5 | 10.2 | 3.9 | 6.9 | 10.5 | 4.5 | 0.8 | 1.4 | 0.5 | 9.0 | 14.0 | 4.2 |
| 3-4 times/week | 2.9 | 5.2 | 1.3 | 3.0 | 5.3 | 1.5 | 0.6 | 0.7 | 0.5 | 4.1 | 7.4 | 1.1 |
| Daily or mostly | 1.4 | 2.4 | 0.8 | 1.7 | 2.5 | 1.1 | 0.1 | 0.2 | 0.0 | 1.6 | 2.9 | 0.2 |
| Missing/unknown | 1.1 | 1.1 | 1.1 | 1.2 | 1.1 | 1.2 | 0.9 | 1.2 | 0.8 | 0.9 | 0.8 | 1.1 |
| **Diabetes (%)** |  |  |  |  |  |  |  |  |  |  |  |  |
| Yes | 8.6 | 9.9 | 7.7 | 5.8 | 6.7 | 5.3 | 11.9 | 12.6 | 11.6 | 16.5 | 18.3 | 14.8 |
| No | 89.4 | 87.8 | 90.6 | 92.3 | 91.1 | 93.1 | 86.0 | 84.3 | 86.9 | 81.3 | 79.4 | 83.1 |
| Missing/unknown | 2 | 2.3 | 1.7 | 1.9 | 2.2 | 1.7 | 2.1 | 3.1 | 1.5 | 2.2 | 2.3 | 2.0 |

\*Values are median (IQR, Q1-Q3). Abbreviations: ALT, Alanine transaminase; BMI, Body mass index; CRP, C-reactive protein; DBP, Diastolic blood pressure; HbA1c, Glycated hemoglobin A1c; HDL-C, High density lipoprotein cholesterol; LDL-C, Low density lipoprotein cholesterol; PBF, Percent body fat; PWV, Pulse wave velocity; SBP, Systolic blood pressure; WC, Waist circumference

Baseline characteristics of participants based on PWV categories in total participants and stratified ethnicity are presented in Table S8 and Table S9, respectively. In the total participants, those with lower PWV were younger, had lower WC, PBF, SBP, DBP, and ALT levels and slightly higher handgrip strength, and they were highly educated, more likely to smoke and drink alcohol, and had a lower prevalence of self-reported diabetes compared to the participants with higher PWV (Table S8). Similar results were observed in men and women (Table S8), and among all three ethnic groups (Table S9).

### EWAS analysis

The mean and SD of 54 predictor variables that were evaluated in relation to PWV are shown in Table S10. Of these variables, 41 variables were significantly associated with PWV at FDR <5% (Figure 1). These comprised positive associations with age (≥60 years, 55-59, 50-54, 45-49, 40-44, 35-39), SBP, HbA1c, insulin, glucose, WC, HC, WHR, PBF, VFL, BFM, FFM, SMM, weight, BMI, height, DBP, triglycerides, TC/HDL-C ratio, ALT, GGT, albumin, CRP, uric acid, ethnicity (Malay, South Asian), and smoking status (former smokers), whereas inverse associations were observed with HDL-C, bilirubin, potassium, vitamin D, educational level, and handgrip strength. The detailed estimates and q values of the evaluated variables for PWV are presented in Table 3 (variables were ordered by smaller to larger q value). Furthermore, similar results were observed in the gender-stratified analysis for both men and women (data not shown).

**Figure 1.**
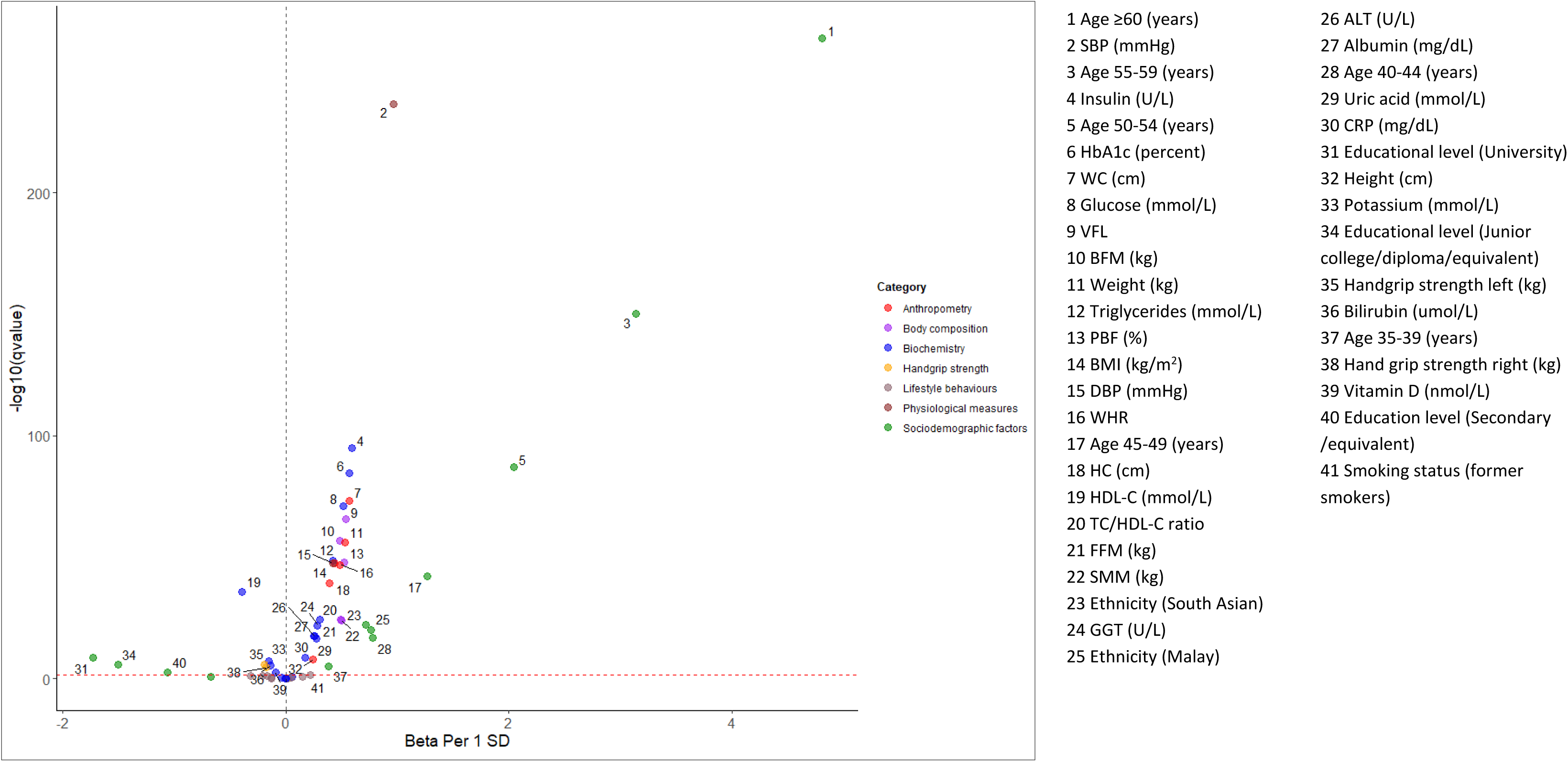
Volcano plot showing associations for PWV with 54 predictor variables in the HELIOS study. The y axis shows −log10 (q value) of the estimated q values from the coefficients of the adjusted linear regression models and the x axis is the estimated beta regression coefficients for one standard deviation change in predictor variable in relation to PWV. The predictor variables are grouped by category by a color. The horizontal red dashed line represents the level of significance corresponding to FDR less than 5%, and the grey dashed line indicates beta of zero with variables to the right of the line indicating a positive association with PWV and those to the left indicating an inverse association with PWV. Abbreviations: ALT, Alanine transaminase; BMI, Body mass index; BFM, Body fat mass; CRP, C-reactive protein; DBP, Diastolic blood pressure; FFM, Fat free mass; GGT, Gamma-glutamyl transferase; HbA1c; Glycated hemoglobin A1c; HDL-C, High-density lipoprotein cholesterol; HC, Hip circumference; PBF, Percent body fat; PWV, Pulse wave velocity; SBP, Systolic blood pressure; SD, standard deviation; SMM, Skeletal muscle mass; TC/HDL-C, Total cholesterol to high-density lipoprotein cholesterol ratio; VFL, Visceral fat level; WC, Waist circumference; WHR, Waist to hip ratio.

**Table 3.**
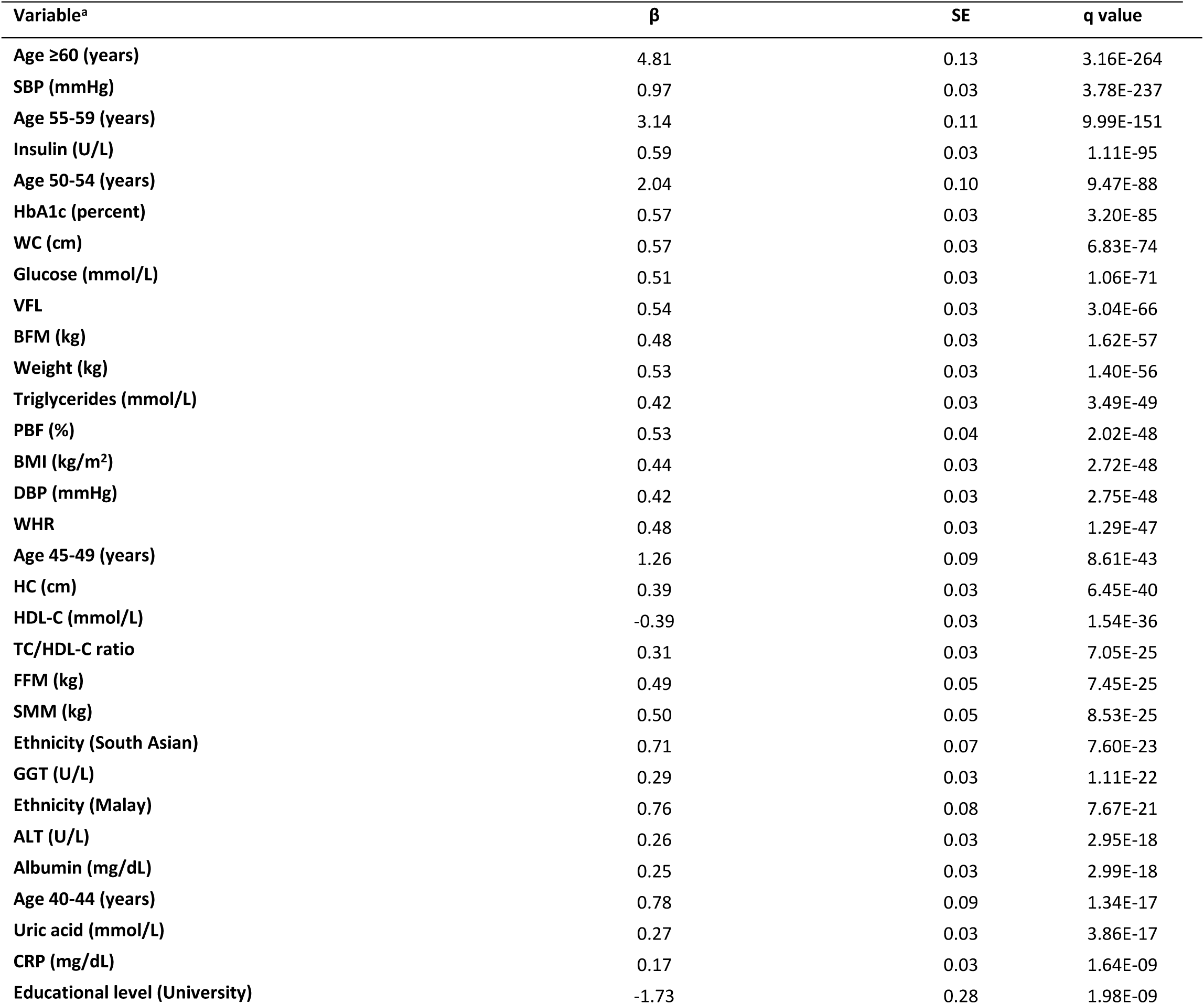
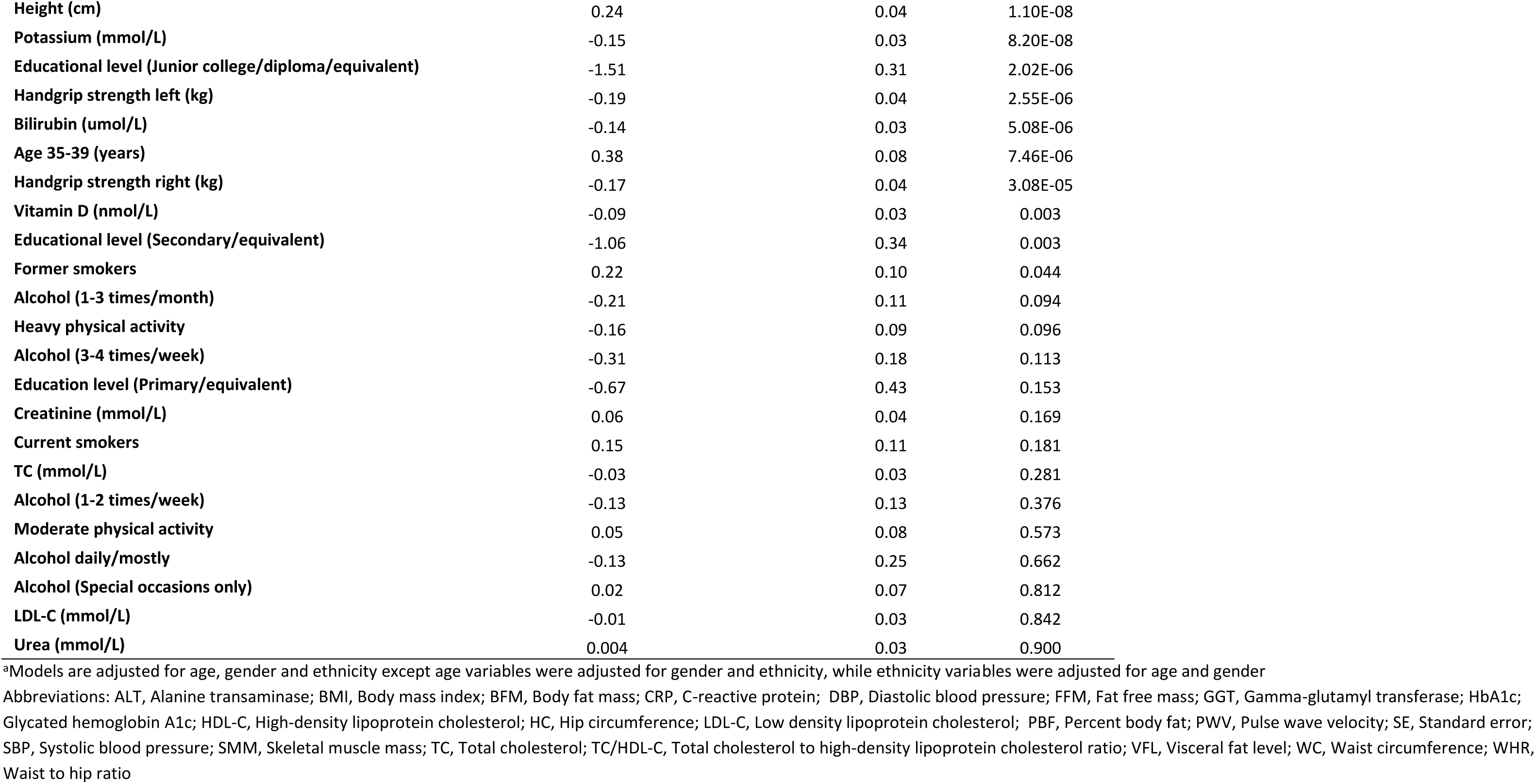
Adjusted beta regression coefficients for PWV with 54 predictor variables in the HELIOS study.

In stratified analysis by ethnicity, we found 39 variables were significantly associated with PWV in Chinese participants (Figure S1 and estimates are presented in Table S11), while 30 variables were significantly associated with PWV in Malay and South Asian participants (Figure S2 and S3, respectively and estimates are presented in Table S11). Exclusion of self-reported diabetes and hypertension subjects did not substantially change the results (Figure S4 and S5 and estimates are presented in Table S12 and S13, respectively).

### Correlations

Evaluation of Pearson correlations showed correlations for several predictor variables (Figure 2). Strong correlations were observed between anthropometric and body composition measurements (r >0.8). Additionally, we found correlations between insulin, triglycerides, HDL-C, anthropometric and body composition measurements (r >0.4) and, as expected, between SBP and DBP (0.76) and TC and LDL-C (0.93).

**Figure 2.**
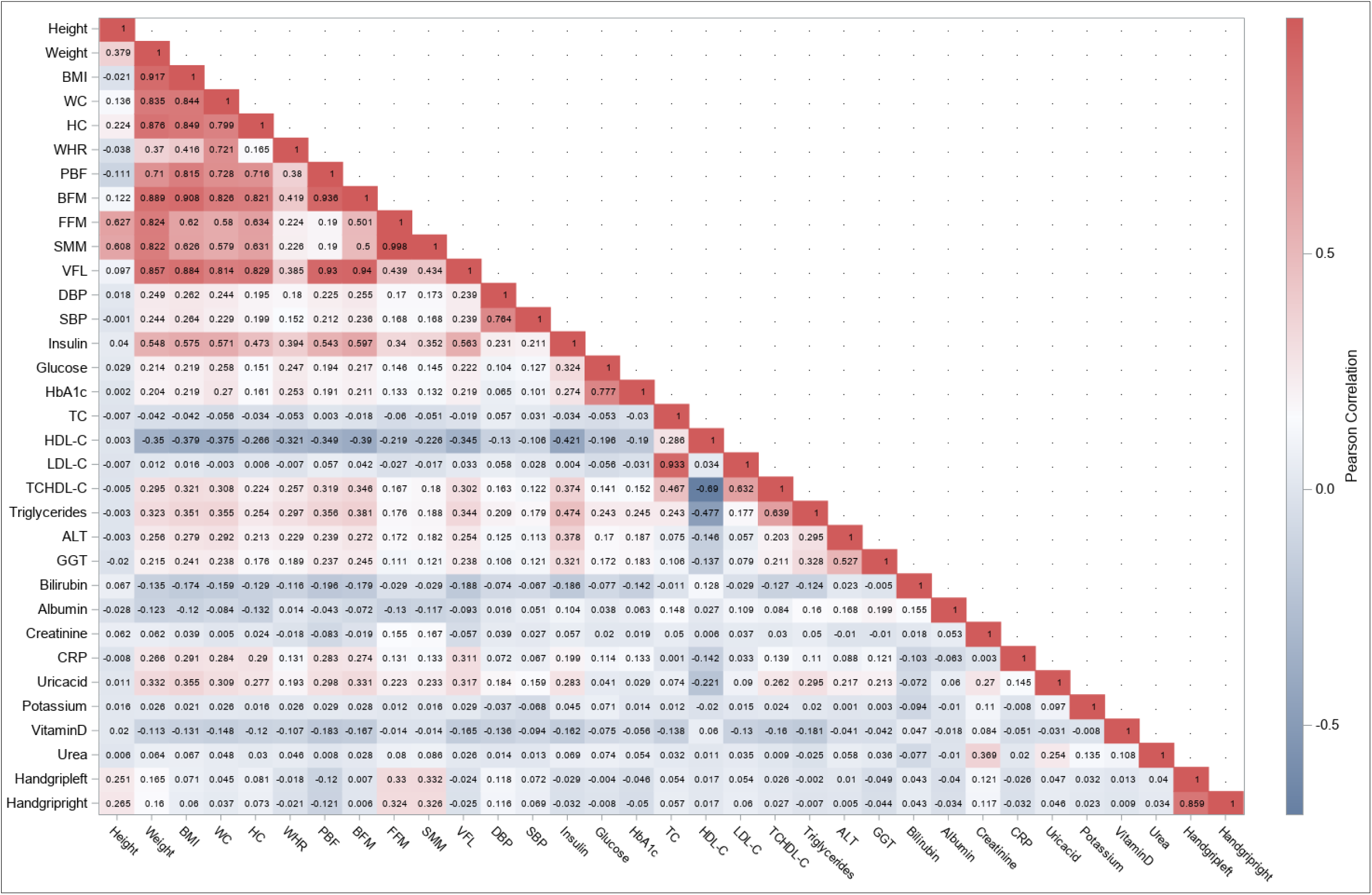
Heat map of Pearson correlation coefficients for the continuous predictor variables in the HELIOS study. Abbreviations: ALT, Alanine transaminase; BMI, Body mass index; BFM, Body fat mass; CRP, C-reactive protein; DBP, Diastolic pressure; FFM, Fat free mass; GGT, Gamma-glutamyl transferase; HbA1c; Glycated hemoglobin A1c; HDL-C, High-density lipoprotein cholesterol; HC, Hip circumference; LDL-C, Low density lipoprotein cholesterol; PBF, Percent body fat; PWV, Pulse wave velocity; SBP, Systolic blood pressure; SMM, Skeletal muscle mass; TC, Total cholesterol; TC/HDL-C, Total cholesterol to high-density lipoprotein cholesterol ratio; VFL, Visceral fat level; WC, Waist circumference; WHR, Waist to hip ratio

### Multivariable analysis

In multivariable analysis, we found 7 variables independently associated with PWV (Table 4). These comprised significant positive associations with HbA1c, SBP, albumin, and inverse associations with HDL-C, potassium, educational level, and handgrip strength (Table 4). In ethnicity-stratified multivariable analysis, HbA1c and SBP were significantly positively associated with PWV across the three ethnic groups (Table 4). In Chinese and Malay participants, albumin was significantly positively associated with PWV, while HDL-C was significantly inversely associated with PWV (Table 4). Furthermore, in Chinese participants, significant inverse associations were observed for PWV with potassium, educational level, and handgrip strength (Table 4).

**Table 4.** Multivariate analysis for PWV by ethnicity in the HELIOS study.

| All<sup>a</sup> |  |  |  | Chinese<sup>b</sup> |  |  |  | Malay<sup>b</sup> |  |  |  | South Asian<sup>b</sup> |  |  |  |
| --- | --- | --- | --- | --- | --- | --- | --- | --- | --- | --- | --- | --- | --- | --- | --- |
| Variable | $\beta$ | SE | q value | Variable | $\beta$ | SE | q value | Variable | $\beta$ | SE | q value | Variable | $\beta$ | SE | q value |
| SBP (mmHg) | 0.83 | 0.04 | 1.63E-15 | SBP (mmHg) | 0.75 | 0.05 | 2.80E-15 | SBP (mmHg) | 0.99 | 0.15 | 1.71E-09 | SBP (mmHg) | 1.11 | 0.11 | 2.8E-15 |
| HbA1c (percent) | 0.38 | 0.05 | 1.63E-15 | HbA1c (percent) | 0.34 | 0.06 | 1.25E-07 | HbA1c (percent) | 0.51 | 0.13 | 0.001 | HbA1c (percent) | 0.34 | 0.09 | 0.001 |
| Albumin (mg/dL) | 0.22 | 0.04 | 1.79E-08 | Albumin (mg/dL) | 0.23 | 0.04 | 3.87E-07 | Albumin (mg/dL) | 0.46 | 0.15 | 0.008 | Albumin (mg/dL) | 0.15 | 0.11 | 0.501 |
| BMI (kg/m<sup>2</sup>) | 0.06 | 0.05 | 0.272 | BMI (kg/m<sup>2</sup>) | 0.10 | 0.06 | 0.140 | BMI (kg/m<sup>2</sup>) | 0.27 | 0.17 | 0.234 | BMI (kg/m<sup>2</sup>) | -0.14 | 0.12 | 0.501 |
| Educational level (University) | -0.81 | 0.26 | 0.006 | Educational level (University) | -0.85 | 0.33 | 0.022 | Educational level (University) | -0.47 | 0.55 | 0.502 | Educational level (University) | -1.12 | 0.87 | 0.501 |
| Potassium (mmol/L) | -0.09 | 0.04 | 0.025 | Potassium (mmol/L) | -0.13 | 0.04 | 0.003 | Potassium (mmol/L) | 0.14 | 0.14 | 0.493 | Potassium (mmol/L) | 0.08 | 0.10 | 0.617 |
| HDL-C (mmol/L) | -0.15 | 0.05 | 0.003 | HDL-C (mmol/L) | -0.12 | 0.05 | 0.032 | HDL-C (mmol/L) | -0.49 | 0.19 | 0.038 | HDL-C (mmol/L) | -0.18 | 0.14 | 0.501 |
| Bilirubin (umol/L) | -0.03 | 0.04 | 0.500 | Bilirubin (umol/L) | -0.06 | 0.04 | 0.212 | Bilirubin (umol/L) | 0.05 | 0.15 | 0.794 | Bilirubin (umol/L) | 0.07 | 0.09 | 0.617 |
| Handgrip strength (right) | -0.14 | 0.05 | 0.010 | Handgrip strength (right) | -0.20 | 0.06 | 0.002 | Handgrip strength (right) | 0.19 | 0.18 | 0.493 | Handgrip strength (right) | 0.00 | 0.13 | 0.985 |
| CRP (mg/dL) | 0.06 | 0.04 | 0.188 | CRP (mg/dL) | 0.04 | 0.04 | 0.444 | CRP (mg/dL) | 0.02 | 0.19 | 0.895 | CRP (mg/dL) | 0.17 | 0.10 | 0.452 |
| Uric acid (mmol/L) | 0.00 | 0.05 | 0.929 | Uric acid (mmol/L) | 0.02 | 0.05 | 0.748 | Uric acid (mmol/L) | -0.23 | 0.17 | 0.348 | Uric acid (mmol/L) | -0.06 | 0.12 | 0.740 |
| Triglycerides (mmol/L) | 0.01 | 0.04 | 0.891 | Triglycerides (mmol/L) | 0.03 | 0.05 | 0.587 | Triglycerides (mmol/L) | -0.06 | 0.18 | 0.794 | Triglycerides (mmol/L) | -0.04 | 0.12 | 0.751 |
| Vitamin D (nmol/L) | 0.08 | 0.04 | 0.056 | Vitamin D (nmol/L) | 0.08 | 0.05 | 0.140 | Vitamin D (nmol/L) | 0.25 | 0.14 | 0.214 | Vitamin D (nmol/L) | 0.05 | 0.08 | 0.625 |
| ALT (U/L) | 0.01 | 0.04 | 0.849 | ALT (U/L) | 0.01 | 0.05 | 0.805 | ALT (U/L) | -0.14 | 0.15 | 0.502 | ALT (U/L) | 0.12 | 0.11 | 0.504 |

<sup>a</sup>Models are adjusted for age, gender and ethnicity<sup>b</sup>Models are adjusted for age, and gender
Abbreviations: ALT, Alanine transaminase; BMI, Body mass index; CRP, C-reactive protein; HbA1c; Glycated hemoglobin A1c; HDL-C, High-density lipoprotein cholesterol; PWV, Pulse wave velocity; SE, Standard error; SBP, Systolic blood pressure

### Two-sample MR analysis

The MR estimates for the 7 variables (SBP, HbA1c, HDL-C, albumin, potassium, education, and handgrip strength), which were independently associated with PWV are presented in Table 5. Based on the main results using the IVW method, we found genetically predicted SBP was positively associated with arterial stiffness (β[95%-CI] = 0.03 [0.001, 0.005], *p* value 0.004), while genetically predicted HbA1c and HDL-C were inversely associated with arterial stiffness (β[95%-CI] = −0.155 [−0.245, −0.065], *p* value 0.001 and β[95%-CI] = −0.033 [−0.056, −0.010], *p* value 0.004), respectively (Table 5). Sensitivity analyses using MR-Egger and weighted median methods yielded similar results for SBP, HDL-C and HbA1c but the associations for HDL-C and HbA1c were not significant (Table 5). No associations were found between genetically predicted albumin, potassium, education, handgrip strength and arterial stiffness (Table 5). The scatter plots of the associations are shown in Figure S6 and S7. In MVMR analysis, the inverse association between genetically predicted HDL-C and arterial stiffness remained after accounting LDL-C and triglycerides (β[95%-CI] = −0.042, *p* value 0.001) (data not shown).

**Table 5.**
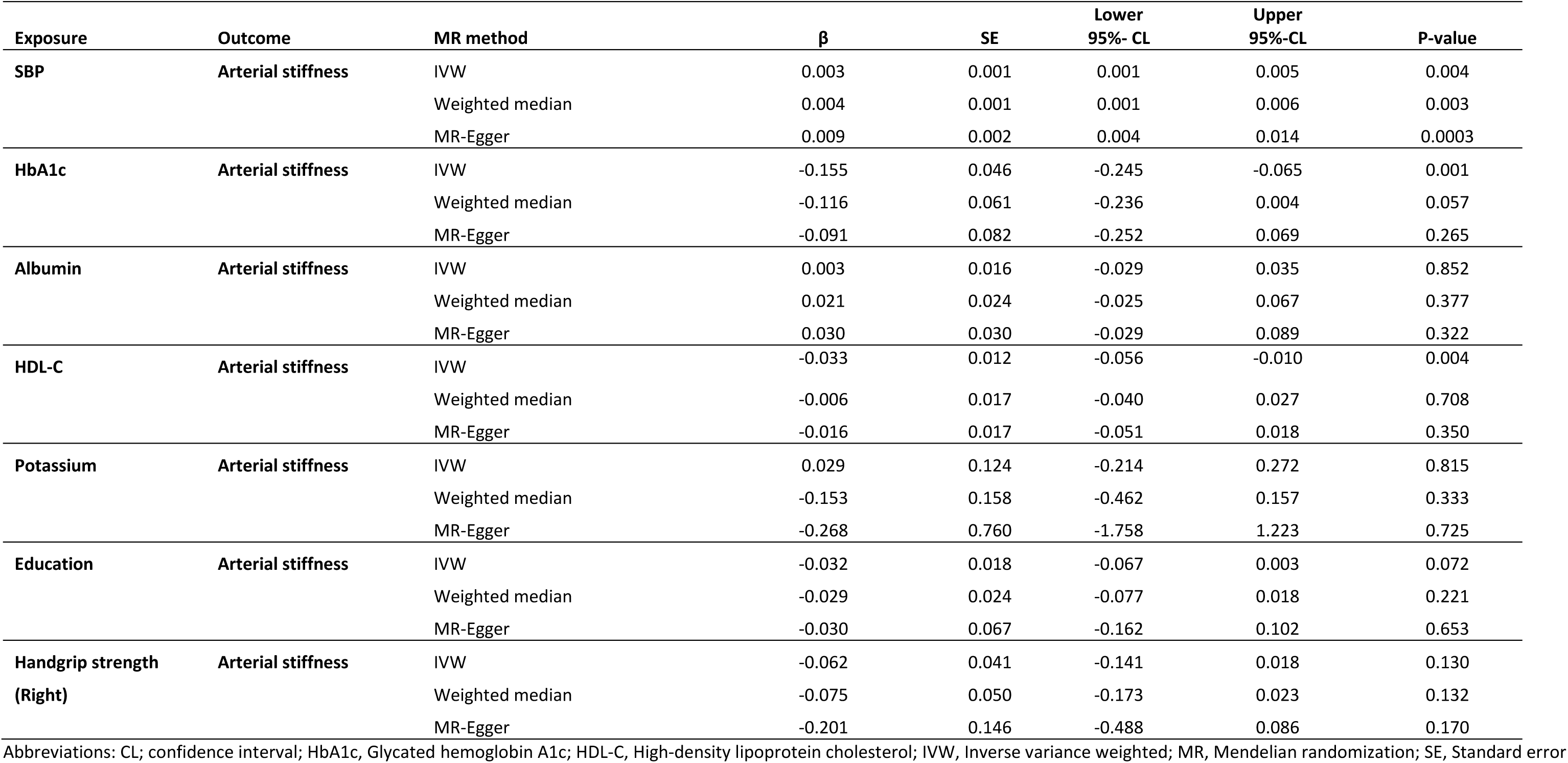
Two sample MR estimates from each method of assessing the causal effect of SBP, HbA1c, albumin, HDL-C, potassium, handgrip strength and education on Arterial stiffness.

| Exposure | Outcome | MR method | $\beta$ | SE | Lower 95%- CL | Upper 95%-CL | P-value |
| --- | --- | --- | --- | --- | --- | --- | --- |
| SBP | Arterial stiffness | IVW | 0.003 | 0.001 | 0.001 | 0.005 | 0.004 |
|  |  | Weighted median | 0.004 | 0.001 | 0.001 | 0.006 | 0.003 |
|  |  | MR-Egger | 0.009 | 0.002 | 0.004 | 0.014 | 0.0003 |
| HbA1c | Arterial stiffness | IVW | -0.155 | 0.046 | -0.245 | -0.065 | 0.001 |
|  |  | Weighted median | -0.116 | 0.061 | -0.236 | 0.004 | 0.057 |
|  |  | MR-Egger | -0.091 | 0.082 | -0.252 | 0.069 | 0.265 |
| Albumin | Arterial stiffness | IVW | 0.003 | 0.016 | -0.029 | 0.035 | 0.852 |
|  |  | Weighted median | 0.021 | 0.024 | -0.025 | 0.067 | 0.377 |
|  |  | MR-Egger | 0.030 | 0.030 | -0.029 | 0.089 | 0.322 |
| HDL-C | Arterial stiffness | IVW | -0.033 | 0.012 | -0.056 | -0.010 | 0.004 |
|  |  | Weighted median | -0.006 | 0.017 | -0.040 | 0.027 | 0.708 |
|  |  | MR-Egger | -0.016 | 0.017 | -0.051 | 0.018 | 0.350 |
| Potassium | Arterial stiffness | IVW | 0.029 | 0.124 | -0.214 | 0.272 | 0.815 |
|  |  | Weighted median | -0.153 | 0.158 | -0.462 | 0.157 | 0.333 |
|  |  | MR-Egger | -0.268 | 0.760 | -1.758 | 1.223 | 0.725 |
| Education | Arterial stiffness | IVW | -0.032 | 0.018 | -0.067 | 0.003 | 0.072 |
|  |  | Weighted median | -0.029 | 0.024 | -0.077 | 0.018 | 0.221 |
|  |  | MR-Egger | -0.030 | 0.067 | -0.162 | 0.102 | 0.653 |
| Handgrip strength<br>(Right) | Arterial stiffness | IVW | -0.062 | 0.041 | -0.141 | 0.018 | 0.130 |
|  |  | Weighted median | -0.075 | 0.050 | -0.173 | 0.023 | 0.132 |
|  |  | MR-Egger | -0.201 | 0.146 | -0.488 | 0.086 | 0.170 |

Abbreviations: CL; confidence interval; HbA1c, Glycated hemoglobin A1c; HDL-C, High-density lipoprotein cholesterol; IVW, Inverse variance weighted; MR, Mendelian randomization; SE, Standard error

## Discussion

In this EWAS approach, we found that age, SBP, DBP, ethnicity, and all anthropometric, and body composition measurements were positively associated with PWV. Furthermore, biochemical parameters such as insulin, HbA1c, glucose, triglycerides, TC/HDL-C ratio, albumin, ALT, GGT, CRP, and uric acid were positively associated with PWV. Conversely, inverse associations were observed for PWV with HDL-C, potassium, vitamin D, bilirubin, handgrip strength and educational level. These associations appear to be stronger in Chinese participants than in Malay and South Asian participants. In the two-sample MR approach, we found a positive association between genetically predicted SBP and arterial stiffness, while genetically predicted HDL-C was inversely associated with arterial stiffness, independent of LDL-C and triglycerides. Furthermore, genetically predicted HbA1c was inversely associated with arterial stiffness, which contrasts with the findings of the present study.

In our study, age was strongly positively associated with PWV. Age is one of most important determinants of arterial stiffness^4^. Our study also found SBP was more strongly positively associated with PWV than DBP. A meta-analysis of prospective studies reported a positive association for arterial stiffness with SBP, while a weaker association with DBP^25^. The biological evidence suggests that aging is associated with structural influences on elastic arteries including collagen formation, degradation of elastin, and increase of vascular smooth muscle cells (VSMCs)^26^, while elevated blood pressure, mainly pulse pressure increases pulsatile aortic wall stress that promotes elastin degradation^26^; hence these structural changes can lead to arterial stiffening.

Our study found significant positive associations for PWV with HbA1c, insulin, and glucose, which is consistent with previous epidemiological studies^6,27,28^. A previous longitudinal study conducted in China showed a positive association between HbA1c and arterial stiffness (Hazard ratio 1.63, 95%-CI: 1.22-2.18 for highest vs lowest quartile)^6^. Some plausible mechanisms may explain the observed associations between HbA1c, glucose, and insulin and arterial stiffness. Chronic hyperglycemia induces proliferation of VSMCs and increases the production of advanced glycation end products and collagen cross-linking, which can lead to stiffening of artery wall^29^. Furthermore, insulin resistance enhances collagen synthesis and increases the expression of numerous genes involved in the inflammatory process; hence these changes can result in arterial stiffening^29^.

In our study, all anthropometric indicators of excess body weight as well as body composition measurements reflecting excess fat were significantly positively associated with PWV. Previous epidemiological studies showed positive associations for arterial stiffness with BMI, WC, WHR and visceral fat area^7,13,30,31^. Mechanistically, excess fat has been related to metabolic disturbances that include higher adipokines levels such as leptin, interleukin-6, tumor necrosis factor-α, which can stimulate vascular inflammation, endothelial dysfunction, and vascular remodeling, leading to arterial stiffness^32^.

Our study found significant positive associations for PWV with triglycerides, and TC/HDL-C ratio, while a significant inverse association with HDL-C. These results are in line with previous epidemiological studies^8,31,33^. Some plausible mechanisms may explain the association between lipids and arterial stiffness. Excess lipid levels bind to the arterial intima and accumulate in the arterial wall, causing oxidative stress and inflammation, which can contribute to arterial stiffening^34^. Conversely, HDL-C plays a key role in removing excess cholesterol and it exhibits anti-inflammatory and antioxidative properties, which could reduce arterial stiffening^34^.

In our study, liver enzymes (ALT and GGT), inflammation marker (CRP), and uric acid were significantly positively associated with PWV. Similar results have been reported in previous epidemiological studies^9–11,35^. Some potential mechanisms underlying the associations between liver enzymes, CRP and uric acid and arterial stiffness. Higher ALT levels have been associated with obesity, dyslipidemia, diabetes, and hypertension^36^, which can contribute to the development of arterial stiffness. Furthermore, elevated GGT, uric acid and CRP levels may enhance oxidative stress and inflammation, which may promote endothelial dysfunction, leading to stiffening of arteries^37–39^.

Our study found a significant inverse association for PWV with bilirubin (per 1 SD increase), potassium, and vitamin D, which is in line with previous cross-sectional studies^40–42^. The mechanisms for beneficial effects of bilirubin, potassium and vitamin D on arterial stiffness are unknown. Experimental studies have demonstrated that bilirubin can enhance endothelial function by reducing oxidative stress via its antioxidant effects^43^. Furthermore, vitamin D can influence endothelial and smooth muscle cells function by exerting antiproliferative effects on VSMCs ^44^, while low potassium intake has been shown to increase vascular calcification and arterial stiffness^45^.

In our study, we found a significant positive association between albumin and PWV. Two cross-sectional studies reported an inverse association between albumin and arterial stiffness^31,46^. The role of albumin in cardiovascular disease and mortality was established with prospective studies and a meta-analysis reporting an increased risk for cardiovascular disease and mortality with lower levels of albumin^47^.

Our study found a significant inverse association between handgrip strength and PWV. Previous epidemiological studies investigating the association between handgrip strength and arterial stiffness have reported inverse associations^48,49^. The mechanisms underlying the association between handgrip strength and arterial stiffness remain unclear. Emerging evidence from previous studies indicates that release of myokines from skeletal muscle may contribute to enhanced endothelial function and vascular health^50^. Furthermore, in our study, higher educational level was inversely associated with PWV. A recent systematic review and meta-analysis reported lower educational level was positively associated with higher PWV^51^.

In our MR analysis, genetically predicted SBP was positively associated with arterial stiffness, which is in line with a previous MR study^12^. Conversely, inverse association was observed between genetically predicted HDL-C and arterial stiffness, independent of LDL-C and triglycerides. To our knowledge, no previous MR studies have investigated the potential causal association between HDL-C and arterial stiffness. Furthermore, we found genetically predicted HbA1c was inversely associated with arterial stiffness, which is inconsistent with the findings of our study. This discrepancy may potentially be explained by distinct polygenic variants that independently affect HbA1c and arterial stiffness through different biological mechanisms^52^.

Strengths of our study include its sample size, multi-ethnic Asian population, and the EWAS approach, where various predictors were assessed in relation to arterial stiffness, while allowing for the multiple comparisons by calculating FDR. Furthermore, we employed MR approach, which could limit the potential for confounding and reverse causation. Our study also has some limitations. The associations reflect cross-sectional analyses, limiting the ability to draw causal inference. Although we propose here a systematic approach that can give a list of PWV correlates with strong statistical support, further examination to assess which among them are most important requires other designs (e.g., causal inference modelling). The HELIOS study participants are of Asian descent, which limits the generalizability of our findings to other ethnicities. In two-sample MR approach, we used publicly available summary statistics from GWAS studies, which may lack certain information about covariates used in the original GWAS data, which could affect the results. Furthermore, potential sample overlap between exposure and outcome samples could lead to bias. Lastly, we were not able to explore linear associations using summary-level datasets.

## Conclusion

Our systematic evaluation provides new knowledge on the complex array of anthropometric, body composition, physiological, biochemical, behavioral, and sociodemographic correlates of PWV. Our MR findings indicate a potential causal association between SBP, HDL-C, HbA1c and arterial stiffness.

## Data Availability

Requests for access to the HELIOS study data should be submitted to

## Acknowledgements

We thank all the HELIOS study participants and HELIOS study team.

## Abbreviations

ALT: Alanine transaminase
BFM: Body fat mass
BMI: Body mass index
CRP: C-reactive protein
DBP: Diastolic blood pressure
EWAS: Exposure wide association study
FDR: False discovery rate
FFM: Fat free mass
GWAS: Genome wide association study
GGT: Gamma-glutamyl transferase
GLGC: Global Lipids Genetics Consortium
HbA1c: Glycated hemoglobin A1c
HC: Hip circumference
HELIOS: Health for Life in Singapore
HDL-C: High-density lipoprotein cholesterol
LDL-C: Low-density lipoprotein cholesterol
ICBP: International Consortium for Blood Pressure
IVW: Inverse variance weighted
MAGIC: Meta-Analyses of Glucose and Insulin-related traits Consortium
MR: Mendelian Randomization
MVMR: Multivariable Mendelian Randomization
PWV: Pulse wave velocity
PBF: Percent body fat
SBP: Systolic blood pressure
SNP: Selected single nucleotide polymorphism
SSGAC: Social Science Genetic Association Consortium
SD: Standard deviation
SMM: Skeletal muscle mass
TC: Total cholesterol
TC/HDL-C: Total cholesterol to high-density lipoprotein cholesterol ratio
UKB: UK Biobank
VFL: Visceral fat level
VSMCs: Vascular smooth muscle cells
WC: Waist circumference
WHR: Waist to hip ratio

